# Evaluation of the number of undiagnosed infected in an outbreak using source of infection measurements

**DOI:** 10.1101/2020.06.09.20126318

**Authors:** Akiva B Melka, Yoram Louzoun

## Abstract

In times of outbreaks, an essential requirement for better monitoring is the evaluation of the number of undiagnosed infected individuals. An accurate estimate of this fraction is crucial for the assessment of the situation and the establishment of protective measures. In most current studies using epidemics models, the total number of infected is either approximated by the number of diagnosed individuals or is dependent on the model parameters and assumptions, which are often debated. We here study the relationship between the fraction of diagnosed infected out of all infected, and the fraction of infected with known contaminator out of all diagnosed infected. We show that those two are approximately the same in exponential models and across most models currently used in the study of epidemics, independently of the model parameters. As an application, we compute an estimate of the effective number of infected by the COVID-19 virus in Israel.

## Introduction

In the absence of a vaccine or efficient treatment, the control of social contacts through large-scale social distancing measures appears to be the most effective means of mitigation in a pandemic^1-5^. Determining the extent of those measures and their stringency requires an accurate evaluation of the total number of infected individuals along with the fraction of those individuals that have not yet been identified^6-8^. Many parameters can influence this evaluation. For instance, when a disease or a virus has a short incubation period and a relatively small spreading rate compared to its detection rate, the fraction of undiagnosed infected is relatively small and the outbreak can be stopped or, at the least, contained, by isolating the infected individuals from the population^9^. In opposite cases, such as in the HIV, SARS, EBOV, or COVID-19 outbreaks, the fraction of undiagnosed infected can be substantial, and spreading can occur through them^10-12^. Modeling has emerged as an important tool in determining the effectiveness of those measures. It enables to gauge the potential for widespread contagion, cope with associated uncertainty, and inform its mitigation^13-15^.

To estimate the total number of infected from observed infected, one needs to determine the Confirmed Cases Fraction *(CCF)*, defined here as the fraction of confirmed (diagnosed) infected out of all infected (both diagnosed and undiagnosed). The reported number of carriers is heavily influenced by sampling biases. this number is usually incomplete due to the lack of testing capacities, and varying testing protocols^16,17^. We here propose that *CCF* can be estimated through the Known Source Fraction *(KSF)*, defined as the fraction of diagnosed individuals with known contaminators. Epidemiological investigations, even on a limited sample of the confirmed infected individuals, can provide the value of KSF and therefore an estimation of *CCF*. In contrast, CCF can only be directly measured through wide scales surveys. Moreover, the total fraction of infected is usually low, requiring very large surveys to obtain accurate estimates of *CCF*.

## Results

Two main types of predictive models were proposed for epidemics: macroscopic models, using aggregated data at the population scale and microscopic models, incorporating distributed information at the individual level^18,19^. Macroscopic models use stochastic processes or ODEs to predict the evolution of the outbreak on a global scale. The simplest and most common model is the SIR model^20^, where the population is divided into three categories: Susceptible (*S*), Infected (*I*), and Removed (*R*) (Fig. 1 upper scheme). N is the total population. In this model, propagation of the virus depends on the infection rate *β* or the number of contacts between susceptible and infected individuals, and the detection rate *γ* that characterizes the time that infected individuals remain contagious. The Removed category can include individuals that survived the virus and are now immune or deceased patients. If stringent confinement is applied, this category can also simply be all diagnosed individuals since they are now removed from the system and can no longer contaminate other individuals. To model *KSF*, we add a category to a stochastic realization of SIR and other models: Controlled (*C*) that represents the individuals among the Removed for whom the contaminator is known. In practice, each time a Susceptible gets infected, an Infected is chosen to be the contaminator and its identity is recorded. When an individual gets diagnosed, we check the identity of its contaminator and if this contaminator has already been diagnosed, we consider that the newly diagnosed individual is added to the Controlled category (see Fig. 1 for a description). We ignored false positives (diagnosed that are not infected) in the current analysis, as their number is consistently small in most epidemics^21,22^. We further discuss false negatives.

**Figure 1.**
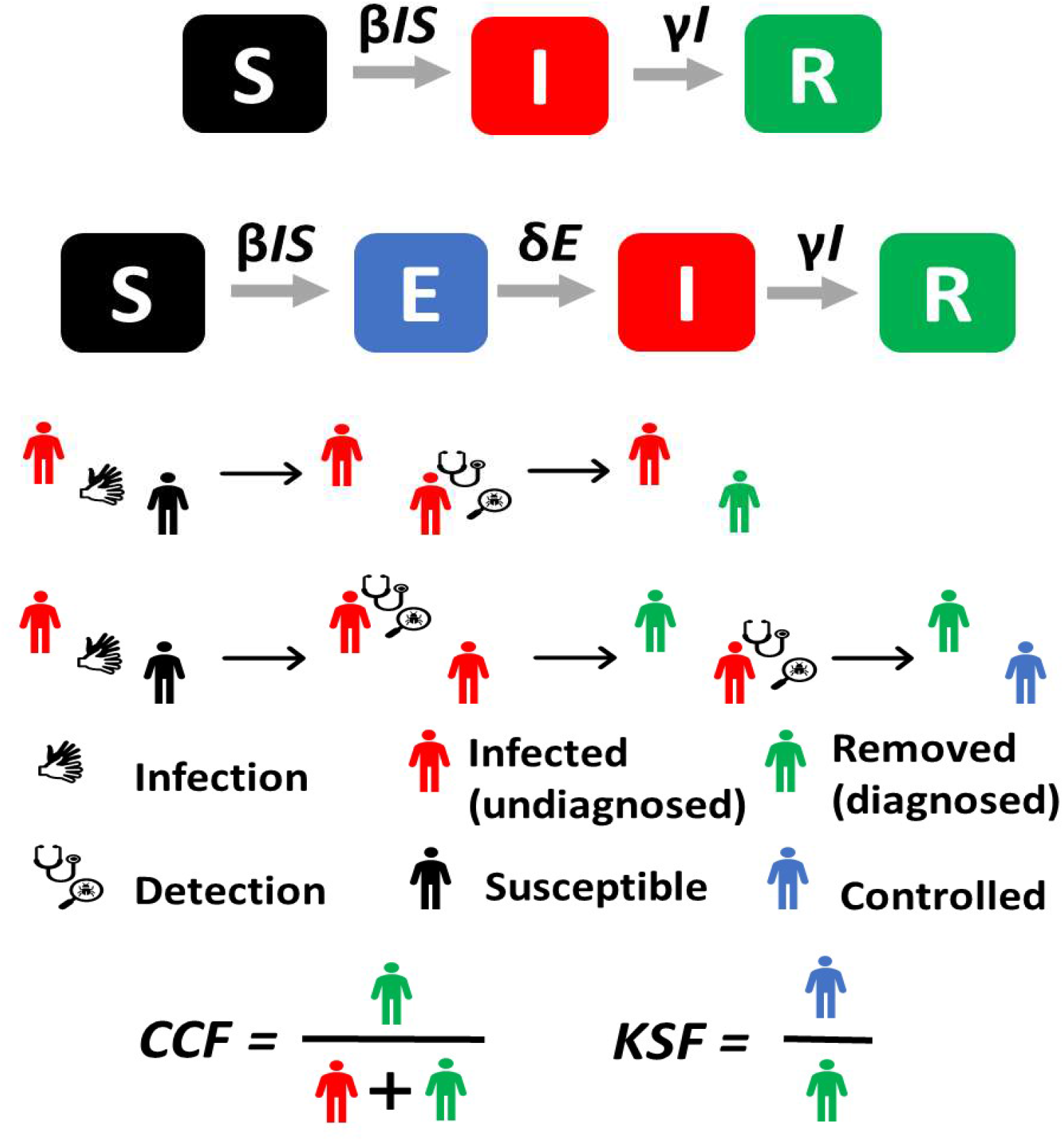
Models Description. (**Upper plot**) dynamics of the SIR model. A Susceptible individual can get infected with a rate proportional to βIS. An infected can get Removed from the system with a rate proportional to γI. (**Middle plot**) dynamics of the SEIR model. An Exposed category is added. Exposed are not infecting but can become infecting with a probability of δ per exposed. (**Lower plot**) dynamics of the discrete-time simulations: First line, each Infected (red) can infect each susceptible (dark). If an infected is detected, it becomes quarantined and thus removed (green). Second line, If the contaminator of a diagnosed individual was already detected (i.e. it is green by the time the new infected is diagnosed), the newly diagnosed is considered controlled (blue) implying that its source of contamination is known. We define two ratios. *CCF* is the fraction of diagnosed individuals over the total number of infected (diagnosed and undiagnosed). *KSF* is the fraction of diagnosed individuals with a known source of contamination.

The first order average dynamics of the SIRC model can be approximated by:

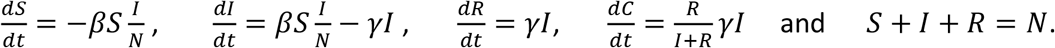

Assuming that initially *S* = *N*, *R* = *C* = 0, and I is very small, and solving for small variations (see Methods for derivations) one gets that *CCF* = *KSF*.

As mentioned above, the evaluation of *CCF* is most relevant when the incubation period is significant. For example, in the current COVID-19 pandemic, a lag of a few days has to be considered when observing infection patterns^23^. Therefore, we also extended the SEIR model to a SEIRC model. In this model, an Exposed (E) category is added that represents the infected individuals that carry the virus but still do not contaminate others (Fig. 1 middle scheme). *δ* is the rate at which an Exposed becomes infectious and can now contaminate others. The dynamics equations are

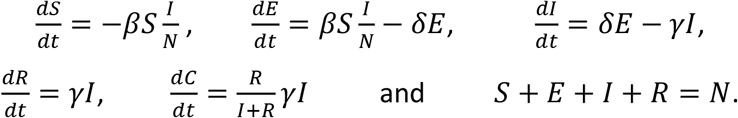

With the same assumptions, we also get that, up to a short and small transient, *CCF* = *KSF*. This can be easily derived from the equations since *KSF* = *C* / *R* and *CCF* = *R* / (*I* + *R*). This equality holds for any model producing an exponential growth of *I*, *R*, and *C*. Moreover, in exponential growth models^14^ (as is observed in real-world at the early stage of most epidemics), we can add that, not only KSF and CCF are equal but also constant (see Methods).

While there is a large number of existing epidemiological models for any pandemic, and specifically for the current COVID-19 pandemic, most current works are based on different versions of either the SIR or SEIR models^23^. More sophisticated versions of SEIR also incorporate migration to assess the efficiency of intercity restrictions^24^, or other categories such as asymptomatic individuals. Finally, models were refined with a time-dependent infection rate, age-dependent infection matrices^25^, or even quarantine.^26,27^. However, the vast majority of these models produce an initial exponential growth, and as such, we expect the equivalence between KSF and CCF to hold.

To validate this equivalence, we tested multiple models. In most realistic cases, the spread dynamics parameters or even the appropriate model are unknown. To show that the relationship between KSF and CCF is not model or parameter specific, we tested this relationship in multiple versions of SIR and SEIR models and different parameter configurations. We implemented SIR and SEIR models with homogenous and heterogeneous infection rates to reflect the fact that not all individuals have the same infection probability (as a function of age/gender/genetics or other factors). In each configuration, we ran simulations with different values of the parameters (Fig. 2c). One can see that, while both fractions vary in different models and parameters, however, an approximately linear relationship is consistent among all models. Besides, those two fractions rapidly achieve equilibrium (Fig. 2a, b).

**Figure 2.**
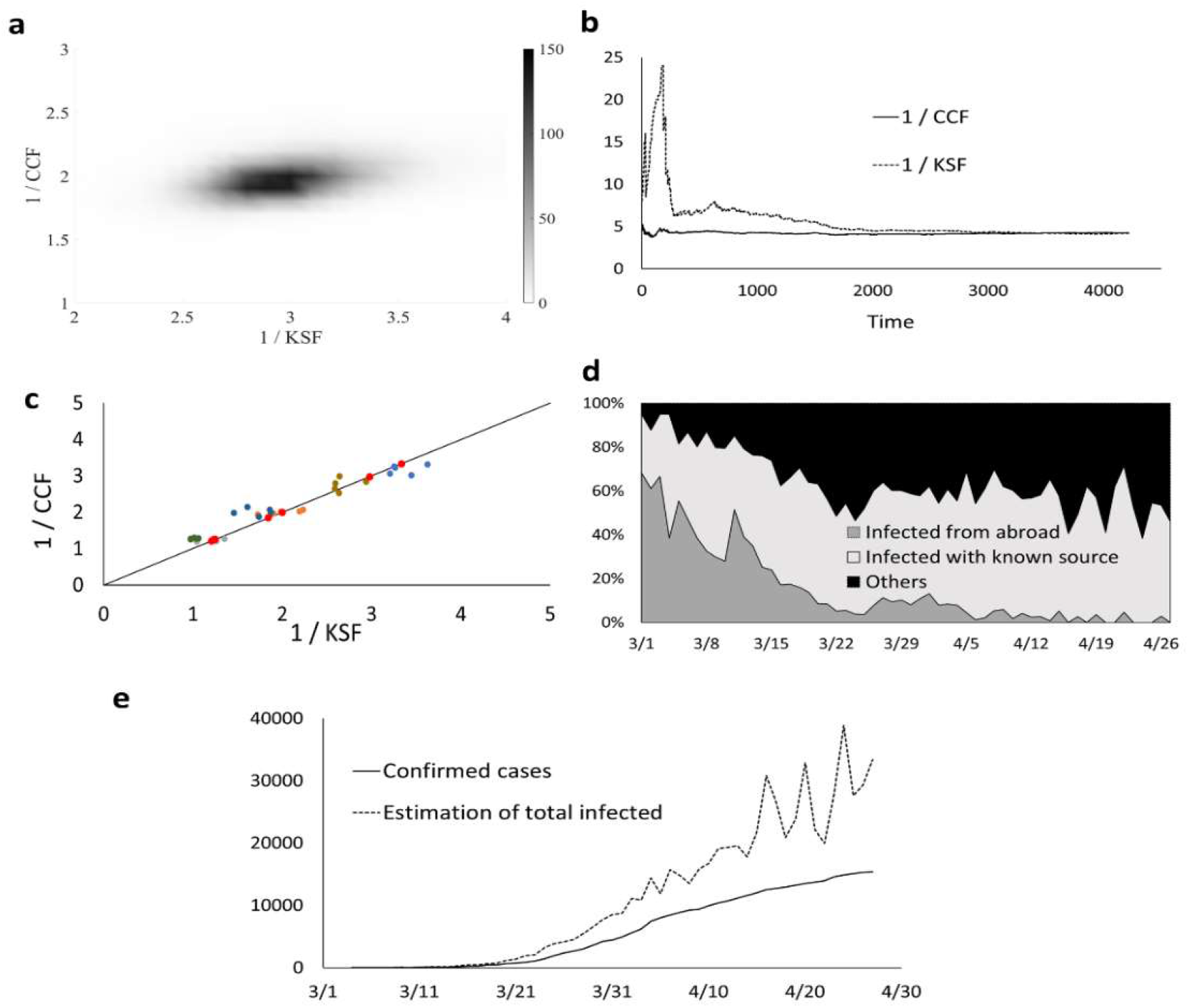
Results from simulations. **(a)** density plot of the two ratios over 1,000 simulations. **(b)** time evolution of the two ratios 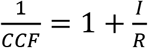 and 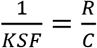. We observe that not only the ratios achieve equilibrium, but they are never very far from it. (c) independently of the model used (SIR or SEIR, with homogeneous or heterogeneous infection rate), we observe a linear relationship between 1/CCF and 1/KSF. (d) distribution of source of infection per day in Israel. Data obtained from the Israeli Ministry of Health with the fraction of confirmed cases with a known source. We ignored in this analysis infected coming from abroad (deep gray). (e) the number of confirmed cases (Removed) in Israel was obtained from world data (full line)^30^. We used our method to estimate the total number of infected in Israel (Dashed line).

Moreover, in different realizations of the same model, most of the trajectory density is centered on a limited range of KSF and CCF values. Different initial conditions and stochastic realizations lead to similar solutions (Fig. 2b). As such, one can use KSF to estimate CCF without further knowledge of the model or its parameters.

At the practical level, since R and C can be obtained from measures of diagnosed infected and epidemiological investigations, KSF can be estimated in most cases. Then CCF and thus I can be determined from the relationship in Fig. 2c. To check the applicability of our methodology and since there is now a large amount of data available, we analyzed the number of confirmed cases for the COVID-19 in Israel every day. In parallel, we analyzed from the Israeli Ministry of Health the fraction of confirmed cases with a known source (KSF) (Fig. 2d). We then estimated the total number of infected in Israel (Fig. 2e).

## Discussion

Multiple models have been proposed to evaluate CCF using, for instance, the number of deceased patients^18^, but in all those studies, the results depend on the models used or on estimates of country-specific parameters, such as the age dependence or the Infection fatality rate. We have presented a method to estimate the fraction of undiagnosed infected from the fraction of infected with a known contaminator (out of all infected). While the first value is hard to measure in realistic situations, the second is often known. The KSF estimate suffers from multiple caveats with opposite effects. First, removed individuals are considered controlled only if their contaminators were already diagnosed when in fact it could be diagnosed even after. Therefore, even already removed individuals could be counted eventually as controlled. A second and more complex problem is that reported infected may be biased toward people who have been in contact with other reported infected. As such, the number of controlled individuals would be overestimated. A direct solution to these limitations would be to perform detailed epidemiological investigations on patients with clinical complications. Such patients typically do not suffer from sampling bias and detailed enough investigations will limit the number of missed controls. Such investigations can be performed on a limited sample^28^. Another limitation of our estimate is that epidemiological investigations are not perfect, as such, some controlled individuals might be missed. Similarly, some diagnosed may be assumed to be infected from a known source, when in fact they were infected by other sources. These limitations can be solved when detailed genetic information is available on the virus or disease. Note again that only a small fraction of the diagnosed individuals needs to be investigated in detail to obtain KSF.

Other versions of the SIR models include a transition to a death state or an Asymptomatic category^29^. Since our Removed category includes all individuals that can no longer contaminate, it already accounts for the dead and the effect of quarantine. Our Infected category includes all undiagnosed individuals that can contaminate others therefore, it accounts for all carriers including the asymptomatic individuals. In the current COVID-19 epidemic, migration has a minor effect on contamination^24^, so we did not include it. However, in the presence of significant migration, the model presented here will not be valid. For the sake of simplicity, we presented here a non-spatial model where all infected individuals can contaminate others disregarding proximity but, since the similarity between CCF and KSF is an inherent property of epidemiological models, we do not expect network and spatial features to change our conclusions.

To summarize, as is the case for every model, multiple caveats can affect the validity of the model, most of those can be avoided in detailed and unbiased investigations on small numbers of diagnosed (even a few tens). Thus, while we do not propose to use the observed relationship as is on biased published epidemiological data, the here reported relationship between KSF and CCF can be a critical tool to estimate the spread of diseases.

## Methods

### SIR Model

The equations for the SIR model can be simplified by using

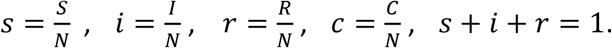

For simplifications, we assume that at inception s = 1, r = c = 0 and compute the first order small variations Δ*_i_*, Δ*_r_*, Δ*_c_*. Δ*_i_*(0) is the initial number of infected individuals.

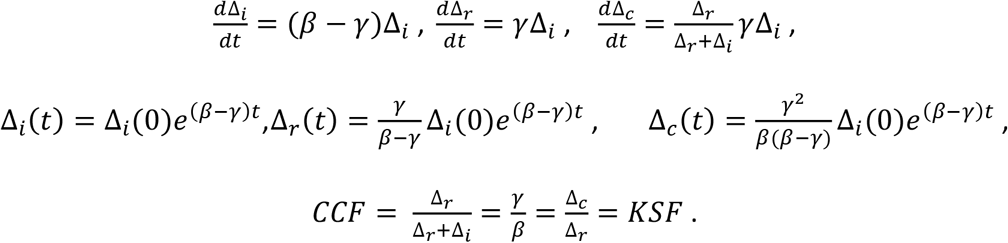

### SEIR model

Using the same notations and assumptions for the SEIR model, one gets

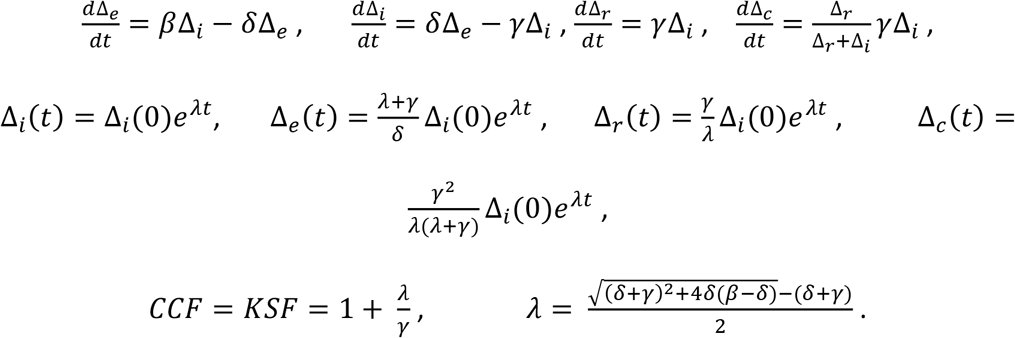

### Simulations

We performed discrete stochastic simulations of both SIR and SEIR models for different infectivity distributions, where each event is explicitly modeled. The models studied either had an equal probability of getting infected for each susceptible, or a variable distribution with a scale-free distribution. We present here results with a slope of −2 in Fig. 2, but other slopes had similar results.

Following is a technical description of the simulation framework. For the sake of efficiency, each event (e.g. infection, detection…) is represented as a tree to allow a rapid selection of the individual involved in the next event. Each leaf corresponds to an individual.

The value of each internal node in the tree is the sum of the values in its direct descendants in the tree. The tree root is the total probability of the event. This configuration enables us to access each individual in logarithmic time. We also keep track of the identity of the contaminator in a repertoire, in case of a contamination event.

We compute the normalized probabilities of each event (based on the top node of the tree of this event) in the appropriate model. At each step, we choose an event based on these probabilities. For a contamination event, a Susceptible is chosen based on its (pre-defined) infectivity. The probability of such an event is the product of the total number of Infected, the total infection probability of Susceptible individuals, and the infection rate β. Following, an infection event, a Susceptible becomes Infected, the chosen Susceptible is determined by traversing the Susceptible tree. The tree is then updated along the entire path. We also randomly choose an Infected as the contaminator and record its leaf number in a repertoire.

For a detection event, an individual is randomly chosen in the Infected tree with a probability proportional to the product of the total number of Infected and the detection rate γ. We then check if his/her contaminator has already been detected by observing if the leaf of the contaminator was already detected. In such a case, the number of Controlled is increased by 1. Once the total number of infected reaches one percent of the total population, we stop the simulation. The ratios in Fig. 2a, b are taken along the simulations. The results in Fig. 2c are at the last time point of the simulation. Simulations where the total number of Infected collapsed before reaching one percent of the total population were not incorporated in the results.

## Data Availability

All data referred to in the manuscript is available.

## Authors contributions

All authors contributed equally.

## Additional information

**Funding:** DoD Grant N629091912097

## References

1 Squazzoni, F., et al. Computational models that matter during a global pandemic outbreak: A call to action. Journal of Artificial Societies and Social Simulation 23 (2020).

2 Prem, K., Liu, Y., Russell, T.W. et al. The effect of control strategies to reduce social mixing on outcomes of the COVID-19 epidemic in Wuhan, China: a modelling study. Lancet Public Health 5, e261-70 (2020).

3 Tian, H., Liu, Y., Li. Y. et al. An investigation of transmission control measures during the first 50 days of the COVID-19 epidemic in China. Science 368, 638—42 (2020).

4 Smith, R. D. Responding to global infectious disease outbreaks: lessons from SARS on the role of risk perception, communication, and management. Social science & medicine 63, 3113–3123 (2006).

5 Punyacharoensin, N., et al. Modelling the HIV epidemic among MSM in the United Kingdom: quantifying the contributions to HIV transmission to better inform prevention initiatives. Aids 29, 339–349 (2015).

6 Glass, K., Becker, N., & Clements, M. Predicting case numbers during infectious disease outbreaks when some cases are undiagnosed. Statistics in Medicine 26, 171–183 (2007).

7 Christaki, E. New technologies in predicting, preventing, and controlling emerging infectious diseases. Virulence 6, 558–565 (2015).

8 Hutchinson, S. J., et al. Method used to identify previously undiagnosed infections in the HIV outbreak at Glenochil prison. Epidemiology & Infection 123, 271–275 (1999).

9 Britton, T., & Scalia Tomba, G. Estimation in emerging epidemics: Biases and remedies. Journal of the Royal Society Interface 16, 20180670 (2019).

10 Yuen, K.S., Ye, Z.W., Fung, S.Y., Chan, C.P. & Jin D.Y, SARS-CoV-2, and COVID-19: The most important research questions. Cell & bioscience 10, 1—5 (2020).

11 Grant, A. Dynamics of COVID-19 epidemics: SEIR models underestimate peak infection rates and overestimate epidemic duration. Preprint at https://www.medrxiv.org/content/medrxiv/early/2020/04/06/2020.04.02.20050674.full.pdf (2020).

12 Richterich, P. Severe underestimation of COVID-19 case numbers: effect of epidemic growth rate and test restrictions. Preprint at https://www.medrxiv.org/content/medrxiv/early/2020/04/17/2020.04.13.20064220.full.pdf (2020).

13 Chowell, G., et al. Model parameters and outbreak control for SARS. Emerging infectious diseases 10, 1258 (2004).

14 Viboud, C., Simonsen, L., & Chowell, G. A generalized-growth model to characterize the early ascending phase of infectious disease outbreaks. Epidemics 15, 27–37 (2016).

15 Finkenstädt, B. F., Bjørnstad, O. N., & Grenfell, B. T. A stochastic model for extinction and recurrence of epidemics: estimation and inference for measles outbreaks. Biostatistics 3, 493–510 (2002).

16 Theagarajan, L.N. Group testing for COVID-19: how to stop worrying and test more. Preprint at https://arxiv.org/pdf/2004.06306.pdf (2020).

17 Flaxman, S., Mishra, S., Gandy, A., et al. Estimating the number of infections and the impact of non-pharmaceutical interventions on COVID-19 in 11 European countries. Preprint at https://arxiv.org/pdf/2004.11342.pdf (2020).

18 Zhigljavsky, A., Whitaker, R., Fesenko, I. et al. Generic probabilistic modelling and non-homogeneity issues for the UK epidemic of COVID-19. Preprint at https://arxiv.org/pdf/2004.01991.pdf (2020).

19 Wearing, H.J., Rohani, P. & Keeling, M.J. Appropriate models for the management of infectious diseases. PLoS medicine 2,174 (2005).

20 Giudici, M., Comunian, A. & Gaburro, R. Inversion of a SIR-based model: a critical analysis about the application to COVID-19 epidemic. Preprint at https://arxiv.org/pdf/2004.07738.pdf (2020).

21 Bai, Y., Yao, L., Wei, T., Tian, F., Jin, D.Y., Chen, L. & Wang, M. Presumed asymptomatic carrier transmission of COVID-19. Jama 323, 1406—7 (2020).

22 Yan, G., Lee, C.K., Lam, L.T. et al. Covert COVID-19 and false-positive dengue serology in Singapore. Lancet Infectious Diseases 20, 536 (2020).

23 Pandey, G., Chaudhary, P., Gupta, R. & Pal, S. SEIR, and Regression Model based COVID-19 outbreak predictions in India. Preprint at https://arxiv.org/ftp/arxiv/papers/2004/2004.00958.pdf (2020).

24 Chinazzi, M., Davis, J.T., Ajelli, M. et al. The effect of travel restrictions on the spread of the 2019 novel coronavirus (COVID-19) outbreak. Science 368, 395—400 (2020).

25 Singh, R. & Adhikari, R. Age-structured impact of social distancing on the COVID-19 epidemic in India. Preprint at https://arxiv.org/pdf/2003.12055.pdf (2020).

26 Berger, D.W., Herkenhoff, K.F. & Mongey, S. An seir infectious disease model with testing and conditional quarantine. National Bureau of Economic Research (2020).

27 Hsieh, Y. H., et al. (2007). Impact of quarantine on the 2003 SARS outbreak: a retrospective modeling study. Journal of Theoretical Biology, 244(4), 729-736.

28 Mueller, M., Derlet, P.M., Mudry, C. & Aeppli, G. Using random testing to manage a safe exit from the COVID-19 lockdown. Preprint at https://arxiv.org/pdf/2004.04614.pdf (2020).

29 Nishiura, H., et al. Estimation of the asymptomatic ratio of novel coronavirus infections (COVID-19). International journal of infectious diseases 94, 154 (2020).

30 https://ourworldindata.org/coronavirus#testing-for-covid-19

